# Comparison of lipidomic profiles sampled with electroporation-based biopsy from healthy skin, squamous cell carcinoma, and basal cell carcinoma

**DOI:** 10.1101/2024.02.01.24301914

**Authors:** Leetal Louie, Julia Wise, Ariel Berl, Ofir Shir-az, Vladimir Kravtsov, Zohar Yakhini, Avshalom Shalom, Alexander Golberg, Edward Vitkin

**Affiliations:** Porter School of Environment and Earth Sciences. Tel Aviv University, Tel Aviv, Israel; Arazi School of Computer Science, Reichman University, Herzliya, Israel; Department of Plastic Surgery, Meir Medical Center, Kfar Sava, Israel; Department of Pathology, Meir Medical Center, Kfar Sava, Israel; Department of Computer Science, Technion - Israel Institute of Technology, Haifa, Israel

**Keywords:** High-throughput lipidomics, electroporation-based biopsy, e-biopsy, cutaneous squamous cell carcinoma, basal cell carcinoma, lipidomic profiles

## Abstract

Incidence rates of cutaneous squamous cell carcinoma (cSCC) and basal cell carcinoma (BCC) are increasing, while the current diagnostic process is time consuming. We introduce a high-throughput sampling approach, termed e-biopsy, utilizing electroporation-based biopsy for efficient collection of tissue lipids. Our study identified 168 lipids using ultra-performance liquid chromatography and tandem mass spectrometry (UPLC-MS-MS). The e-biopsy technique demonstrated its ability to profile the human skin lipidome. Comparative analysis revealed 27 differentially expressed lipids (p<0.05). The observed trend of lipidomic profiles was low diglycerides in cSCC and BCC, elevated phospholipids in BCC, and increased lyso-phospholipids in cSCC compared to healthy skin samples. These findings contribute to understanding skin cancers and highlight the potential of e-biopsy for lipidomic analysis in skin tissues.

## 1. Introduction

Rising trends in keratinocyte carcinoma (cSCC and BCC) incidence have been observed[1–5]. These are among the most common cancers diagnosed, but due to their low mortality rate, are usually excluded from cancer registries[4], despite their impact on quality of life and risks of premature mortality[2,6]. These trends foreshadow increasing wait time for diagnosis under the current gold standard diagnostic method which relies on excision and histopathological examination, with advanced technologies acting as support[7]. The current gold standard is problematic when subsequent treatment requires electrodesiccation and cautery[8], so finding less invasive methods for differentiating between healthy and cancerous skin, as well as between cancer types will prove useful in light of incidence trends. Methods that can provide rapid results will also be beneficial considering the difference in aggressiveness and likelihood of metastasis of cSCC and BCC[9].

Collecting samples of molecular information from cSCC and BCC lesioned skin with modern technology like e-biopsy and others[10,11] is simpler and faster than current biopsy methods, alluding to the potential of molecular profiling in diagnostics. cSCC and BCC are cancers of keratinocytes which functionally make and secrete lipids[12], therefore exploring differential expression of their lipid profiles may reveal trends that differentiate the two from each other as well as from healthy skin.

Molecular profiling technology gives us snapshots of cellular fabric where we can observe trends that have physical manifestations. We have entered the era of personalized medicine where biological profiling provides useful information in cancer research and treatment[13–15]. Previously, high-throughput analyses of genes, proteins, and metabolites have been reported for cSCC and BCC[16– 20]. Lipids profiles, however, have mostly been reported for BCC[21–23] using low-throughput methods and serum samples with the exception of one high-throughput cSCC study that included 70 lipids among metabolites[24]. Beyond medicine, molecular profiling plays an important role in other aspects of life such use in agriculture food and safety, forensic science, and environmental monitoring.

The recent electroporation-based biopsy sampling technique, e-biopsy, that leverages cell permeabilization caused by electric fields for molecular harvesting, demonstrated its ability to sample molecular profiles that differentiate between cancerous and healthy skin[25–27]. Coupling this sampling method with UPLC-MS-MS, which has proven its ability to identify potential markers of skin cancer[18], provides a promising direction for high-throughput analysis of extractable molecules.

Here we perform a comparative analysis of the first high-throughput lipidomic profiling of cSCC, BCC, and healthy skin tissue using e-biopsy. These profiles were collected with the e-biopsy approach, thus adding to previous e-biopsy enabled profiling in transcriptomics, proteomics, and metabolomics[16,25–27].

## 2. Material and Methods

### 2.1. Human patients

A list of patient age, sex, and tumor type is provided in **Table 1**. This study was approved by the Meir Medical Center IRB, number MMC-19-0230. All patients gave consent for participation and for performance of genetic analysis of their sample tissue.

**Table 1.**
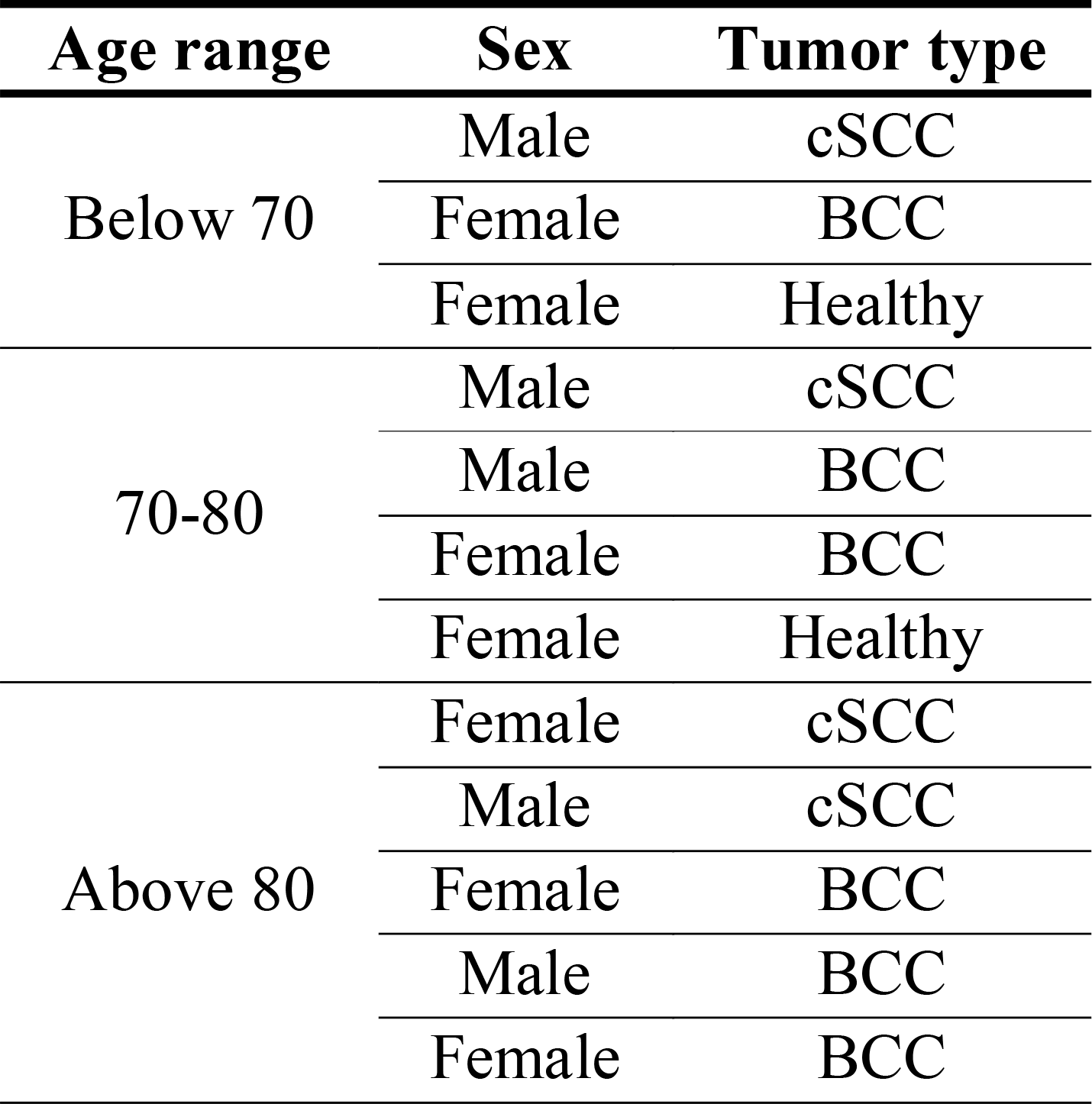
Patient sex, age, and tumor type.

### 2.2. Sample collection

From March 2020 to March 2022, 10 tissue samples were collected from 10 patients who underwent surgical excision of a skin lesion suspected as BCC or cSCC at Meir Medical Center, Israel. Three healthy tissue samples were collected from 2 patients undergoing blepharoplasty. Excised samples were at least 1 cm in diameter. E-biopsy extraction was performed on 13 fresh (between 10-20 minutes after surgery) samples. Lipid analysis was performed via UPLC-MS-MS. **Fig.1** summarizes the workflow and e-biopsy method.

E-biopsy was performed with a custom-made high-voltage pulsed electric field generator under conditions previously described for protein extraction from cSCC and BCC[16]. The liquids sampled were immediately transferred to 1.5ml tubes with 100µl double distilled water and stored at -20 □ until shipped to Beijing Genomics Institute for analysis.

**Figure 1.**
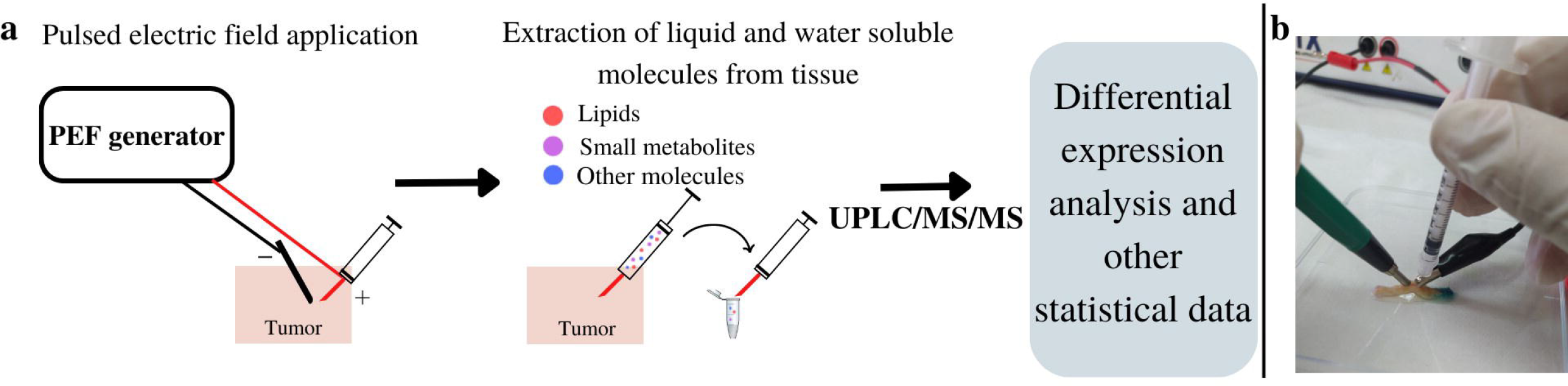
**a-b**. E-biopsy analysis and workflow. **a**. Sample collection through pulsed electric field (PEF) application leads to extraction of water-soluble compounds that underwent subsequent UPLC-MS-MS and differential expression analysis. **b**. Needle and electrode positioning on skin during molecular harvesting by e-biopsy.

### 2.3. UPLC-MS-MS Analysis

The UPLC-MS-MS analysis was performed by Beijing Genomics Institute. An ACQUITY UPLC CSH C18(1.7 μm,2.1*100 mm, Waters, USA) and Q Exactive mass spectrometer (Thermo Fisher Scientific, USA) were used for lipid analysis. The output of the UPLC-MS/MS analysis was imported to LipidSearch v.4.1 software (Thermo Fisher Scientific, USA) for molecular identification and quantification. The software was also used to impute missing values. Excel files were generated to include, among other things, the lipid ID, reliability score (graded A to D, with A and B being the most accurately identified lipids used for subsequent differential lipid screening), and the observed intensity of the lipid in the sample (**Table S1** and **GitHub:** https://github.com/GolbergLab/BCC_SCC_Lipidomics). This data was used for analyses of differential lipid abundance. Detailed UPLC-MS/MS information and methods can be found in the **Supplementary methods**.

### 2.4. Differential lipid screening

The measured lipid intensities (**Table S1**) were used for differential lipid screening. Student’s T-test, and fold change between average measured lipid intensities were calculated for each comparison pair (cSCC vs. Healthy, BCC vs. Healthy, cSCC vs. BCC). The resulting p-values and fold change values were used in overabundance analysis and to generate a volcano plot (**Fig. 2 and Fig. 3**).

**Figure 2.**
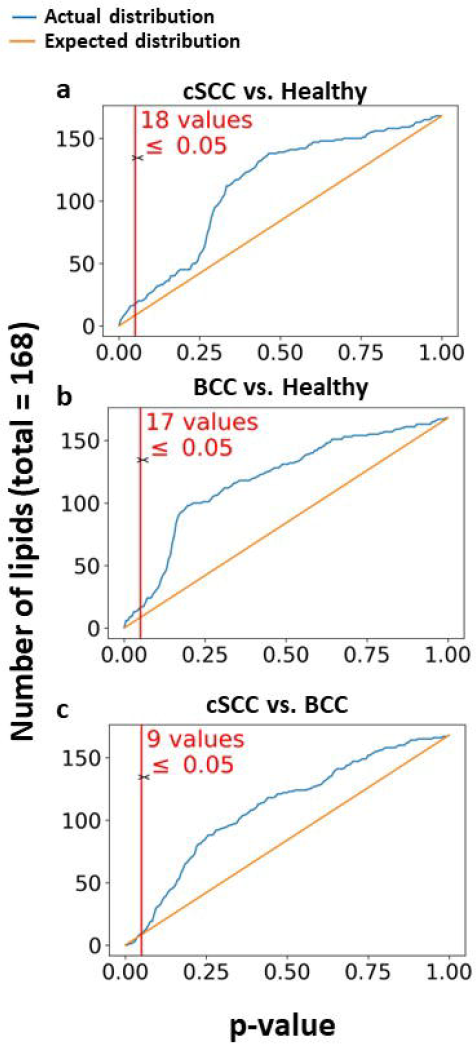
**a-c**. Overabundance plots comparing the distribution of lipid differential expression (both over- and under-expression) p-values between control (normal skin tissue), BCC, and cSCC tumor samples. Total 13 samples, and 168 lipids extracted by e-biopsy were analyzed. Vertical red line marks Student’s T-test pvalue cut-off of 0.05. **a**. cSCC vs. healthy **b**. BCC vs. healthy and **c**. cSCC vs. BCC.

**Figure 3.**
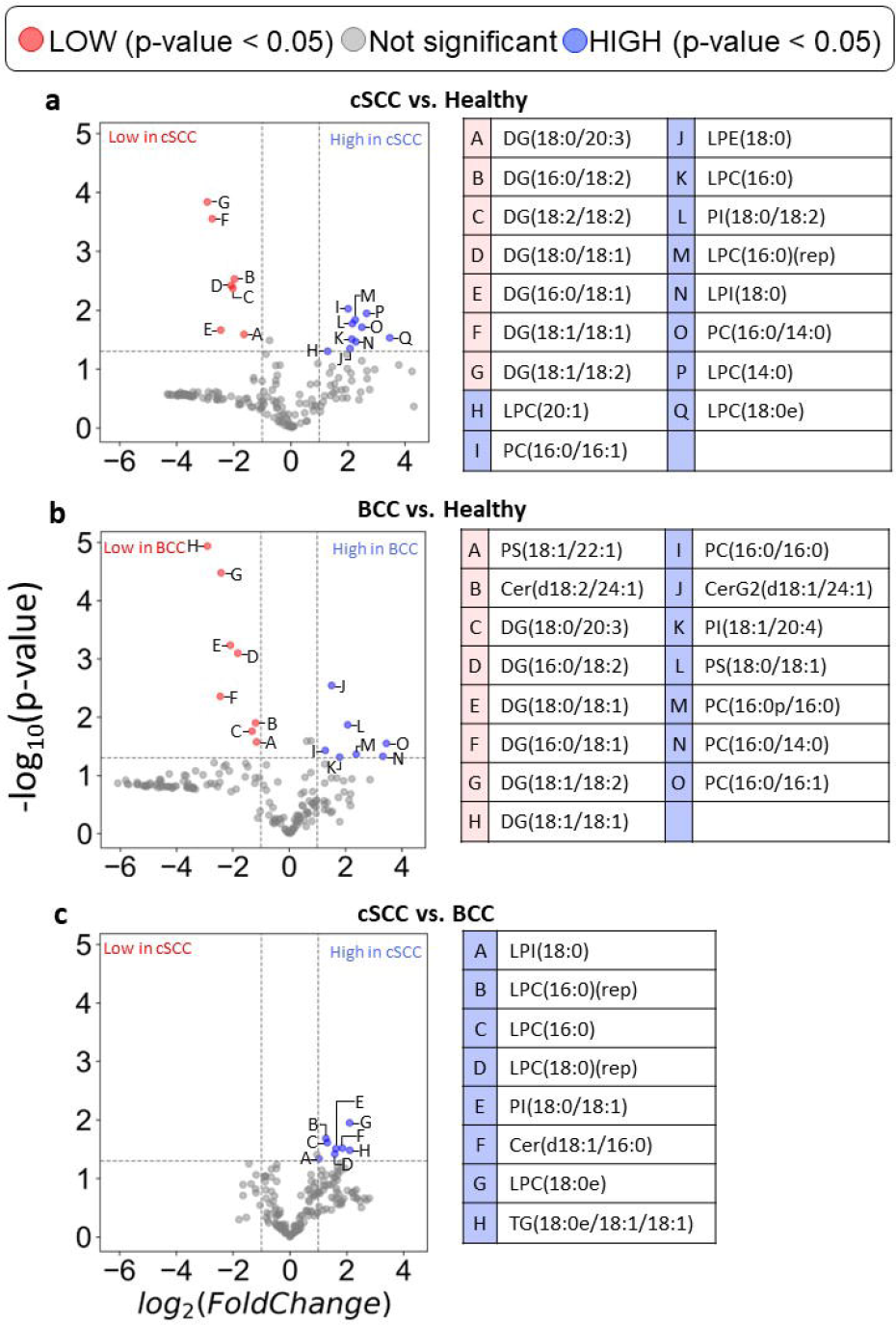
**a-c**. Volcano plots and tables showing the fold change difference of lipid intensities. **a**. cSCC vs. Healthy. **b**. BCC vs. Healthy. **c**. cSCC vs. BCC. Tables with lipid names correspond to lettered data points in the adjacent volcano plots. Fold change and p-value data for lipids listed in the tables can be found in **Table 2, Table S2, Table S3**, and **Table S4**.

#### 2.4.1. Statistical overabundance analysis

Overabundance analysis compares actual and expected distributions of p-values to verify that compounds have different abundance levels when comparing to classes of samples[28]. This approach explores internal data variability and helps addressing multiple comparisons. The analysis relies only on the number of compounds (i.e., lipids) and their observed corresponding p-values (here obtained from the Student’s T-test). The distribution of the expected p-values was generated from a null model assuming the same number of compounds (**Fig. 2**).

**Table 2.** Differentially expressed lipids with associated p-values and fold change values. Columns contain the lipids (in bold) identified as significant by the pairwise comparison of groups: cSCC vs. Healthy, BCC vs. Healthy, cSCC vs. BCC. Also categorized are the lipids found in combined cSCC and BCC groups compared to Healthy, showing the difference between keratinocyte carcinoma and healthy skin. Lastly, lipids of cSCC are isolated with results of common lipids that differentiate cSCC from BCC and healthy skin. Reading example: Cer(d18:2/24:1) has Student’s T-test p-value of 0.01 and the ratio of its average intensity in BCC tissues to its average intensity in healthy tissues is 0.44.

|  | **cSCC vs. Healthy** | **BCC vs. Healthy** | **cSCC<sup>SH</sup> and BCC<sup>BH</sup> vs. Healthy** | **cSCC vs. BCC** | **cSCC vs. BCC<sup>SB</sup> and Healthy<sup>SH</sup>** |
| --- | --- | --- | --- | --- | --- |
|  | **DG(18:2/18:2)** | **Cer(d18:2/24:1)** | **DG(16:0/18:1)** | **Cer(d18:1/16:0)** | **LPC(16:0)** |
| P-value | 0.004 | 0.01 | 0.02<sup>SH</sup>, 0.004<sup>BH</sup> | 0.03 | 0.02<sup>SB</sup>, 0.03<sup>SH</sup> |
| Fold Change | 0.25 | 0.44 | 0.18<sup>SH</sup>, 0.18<sup>BH</sup> | 3.59 | 2.4883<sup>SB</sup>, 4.4<sup>SH</sup> |
|  | **LPC(14:0)** | **CerG2(d18:1/24:1)** | **DG(16:0/18:2)** | **LPC(18:0)(rep)** | **LPC(16:0)(rep)** |
| P-value | 0.01 | 0.003 | 0.003<sup>SH</sup>, 0.0008<sup>BH</sup> | 0.04 | 0.02<sup>SB</sup>, 0.01<sup>SH</sup> |
| Fold Change | 6.3 | 2.84 | 0.26<sup>SH</sup>, 0.28<sup>BH</sup> | 2.98 | 2.41<sup>SB</sup>, 4.77<sup>SH</sup> |
|  | **LPC(20:1)** | **PC(16:0/16:0)** | **DG(18:0/18:1)** | **PI(18:0/18:1)** | **LPC(18:0e)** |
| P-value | 0.05 | 0.04 | 0.004<sup>SH</sup>, 0.0006<sup>BH</sup> | 0.03 | 0.01<sup>SB</sup>, 0.03<sup>SH</sup> |
| Fold Change | 2.45 | 2.43 | 0.24<sup>†</sup>, 0.24<sup>BH</sup> | 3.07 | 4.29<sup>SB</sup>, 11.07<sup>SH</sup> |
|  | **LPE(18:0)** | **PC(16:0p/16:0)** | **DG(18:0/20:3)** | **TG(18:0e/18:1/18:1)** | **LPI(18:0)** |
| P-value | 0.05 | 0.04 | 0.03<sup>SH</sup>, 0.02<sup>BH</sup> | 0.03 | 0.05<sup>SB</sup>, 0.03<sup>SH</sup> |
| Fold Change | 4.21 | 5.21 | 0.32<sup>SH</sup>, 0.40<sup>BH</sup> | 4.3 | 2.03<sup>SB</sup>, 4.86<sup>SH</sup> |
|  | **PI(18:0/18:2)** | **PI(18:1/20:4)** | **DG(18:1/18:1)** |  |  |
| P-value | 0.02 | 0.05 | 0.0003<sup>SH</sup>, 1.16E-05<sup>BH</sup> |  |  |
| Fold Change | 4.46 | 3.46 | 0.15<sup>SH</sup>, 0.13<sup>BH</sup> |  |  |
|  |  | **PS(18:0/18:1)** | **DG(18:1/18:2)** |  |  |
| P-value |  | 0.01 | 0.0001<sup>SH</sup>, 3.33E-05<sup>BH</sup> |  |  |
| Fold Change |  | 4.22 | 0.13<sup>SH</sup>, 0.19<sup>BH</sup> |  |  |
|  |  | **PS(18:1/22:1)** | **PC(16:0/14:0)** |  |  |
| P-value |  | 0.03 | 0.02<sup>SH</sup>, 0.05<sup>BH</sup> |  |  |
| Fold Change |  | 0.45 | 5.61<sup>SH</sup>, 10.02<sup>BH</sup> |  |  |
| Summary of lipids | Diglyceride<br>Lyso-phospholipids<br>Phospholipid | Ceramides<br>Phospholipids | Diglyceride<br>Phospholipid | Ceramides<br>Lyso-phospholipids<br>Phospholipids<br>Triglycerides | Lyso-phospholipids |
| DG Diglyceride |  |  |  |  |  |
| Cer Ceramides |  |  |  |  |  |
| CerG2 Diglycosylceramide |  |  |  |  |  |
| LPC Lyso-phosphatidylcholine |  |  |  |  |  |
| LPE Lyso-phosphatidylethanolamine |  |  |  |  |  |
|  |  |  | LPI Lyso-phosphatidylinositol |  |  |
|  |  |  | PI Phosphatidylinositol |  |  |
|  |  |  | PC Phosphatidylcholine |  |  |
|  |  |  | PS Phosphatidylserine |  |  |
|  |  |  | TG Triglyceride |  |  |

<sup>†</sup>(rep) refers to repeated lipid identification results

#### 2.4.2. Volcano plot analysis

Volcano plot overlays the magnitude of fold change of lipids with their differential significance between two analyzed populations. Differentially expressed lipids were defined for this analysis as those with a *-log10(p-value)>1*.*3* (i.e. *p-value<0*.*05*) and those with *-1<log2(fold-change)<1*. Fold change calculations were carried out using the average of intensity values for each comparison group i.e., *fold-change(lipid) = avg(grp1) / avg(grp2)*. The data was then filtered to include only the compounds with high reliability score (grades A and B) (**Fig. 3)**. The data of the most interesting compounds were compiled into tables to showcase their associated p-values and fold change values (**Table 2**).

## 3. Results

The initial 309 identified lipids were filtered according to reliability score, omitting lipids with grades C and D, which resulted in 168 lipids eligible for differential expression analysis. The analysis was performed for each of three comparison configurations: cSCC vs. Healthy, BCC vs. Healthy, and cSCC vs. BCC. Overabundance plots (**Fig. 2**) represent analysis results, where the number of lipids with Student’s T-test p-value below 0.05 is highlighted in red. Volcano plots (**Fig. 3**) show relative group affinity of each lipid, highlighting significantly over- and under-expressed lipids. The major findings are summarized in **Table 2**.

### 3.1. Electroporation sampled lipidomic profile differentiate cSCC vs. Healthy skin

The overabundance analysis of cSCC compared to Healthy showed a total of 18 lipids with Student’s T-test p-values below 0.05 (**Fig. 2a**, corresponding to FDR=0.47). Moreover, 6 of these lipids resulted in p-value below 1e-2 (FDR=0.28) and 2 of them in p-value below 1e-3 (FDR=0.08). Of these, 7 were significantly lower in cSCC and 10 significantly higher in cSCC (**Fig. 3a)**. All observed under-expressed cSCC lipids were diglycerides, specifically (in increasing negative fold change): DG(18:0/20:3), DG(16:0/18:2), DG(18:2/18:2), DG(18:0/18:1), DG(16:0/18:1), DG(18:1/18:1), and DG(18:1/18:2) (**Fig. 3a**). Lipids with higher expression in cSCC were phospholipids and lyso-phospholipids, specifically (in increasing positive fold change): LPC(20:1), PC(16:0/16:1), LPE(18:0), LPC(16:0), PI(18:0/18:2), LPC(16:0)(rep), LPI(18:0), PC(16:0/14:0), LPC(14:0), and LPC(18:0e) (**Fig. 3a)**. High-resolution volcano plot of this comparison can be viewed in **Fig. S1**. All associated p-values and fold change values for the lipids listed are reported in **Table S2**.

### 3.2. Electroporation sampled lipidomic profile differentiate BCC vs. Healthy skin

The overabundance analysis of BCC compared to Healthy showed a total of 17 lipids with Student’s T-test p-values less than 0.05 (**Fig. 2b**, corresponding to FDR=0.49). Moreover, 6 of these lipids resulted in p-value below 1e-2 (FDR=0.28) and 4 of them in p-value below 1e-3 (FDR=0.04). Of these, 8 were significantly lower in BCC and 7 significantly higher in BCC (**Fig. 3b**). Observed under-expressed BCC lipids were mostly diglycerides as well as a ceramide and phospholipid, specifically (in increasing negative fold change): PS(18:1/22:1), Cer(d18:2/24:1), DG(18:0/20:3), DG(16:0/18:2), DG(18:0/18:1), DG(18:1/18:2), DG(16:0/18:1), and DG(18:1/18:1) (**Fig. 3b**). Lipids with higher expression in BCC were mostly phospholipids and a ceramide, specifically (in increasing positive fold change): PC(16:0/16:0), CerG2(d18:1/24:1), PI(18:1/20:4), PS(18:0/18:1), PC(16:0p/16:0), PC(16:0/14:0), and PC(16:0/16:1) (**Fig. 3b**). Higher levels of phospholipids in BCC versus healthy skin are consistent with previous reports[22]. In contrast to previous reports that found significantly higher TGs in BCC compared to healthy skin[22], all the 65 TGs identified by this study were (not-significantly) lower in BCC compared to healthy tissues. High-resolution volcano plot of this comparison can be viewed in **Fig. S2**. All associated p-values and fold change values for the lipids listed are reported in **Table S3**.

### 3.3. Electroporation sampled lipidomic profile differentiate cSCC vs. BCC skin

The overabundance analysis of cSCC compared to BCC showed a total of 9 lipids with Student’s T-test p-values less than 0.05 (**Fig. 2c**, corresponding to FDR=0.93), with 8 of them significantly higher in cSCC and none significantly lower (**Fig. 3c**). Lipids with higher expression in cSCC compared to BCC were mostly lyso-phospholipids as well as a phospholipid, ceramide, and triglyceride, specifically (in increasing order of fold change): LPI(18:0), LPC(16:0)(rep), LPC(16:0), LPC(18:0)(rep), PI(18:0/18:1), Cer(d18:1/16:0), LPC(18:0e), and TG(18:0e/18:1/18:1) (**Fig. 3c**).

High-resolution volcano plot of this comparison can be viewed in **Fig. S3**. All associated p-values and fold change values for the lipids listed are reported in **Table S4**.

## 4. Discussion

We performed a comparison of high-throughput lipidomic profiles sampled with e-biopsy from healthy, cSCC, and BCC skin tissues. The overabundance and volcano plot analyses suggest a difference in lipidomic profiles between cancer and healthy skins, with a general trend of lower DGs and higher phospholipid subclasses in cancerous tissue. There was also a slight difference and a separation potential between cSCC and BCC lipid profiles as higher intensities of phospholipids and other lipids were observed in cSCC. The comparison of cSCC to healthy tissue revealed lower DGs and higher phospholipids and lyso-phospholipids. Similarly, the comparison of BCC to healthy skin found lower diglycerides and higher phospholipids. In the BCC to healthy tissue comparison, 2 ceramides were identified, 1 higher and the other lower in BCC. In the comparison of cSCC to BCC, several lyso-phospholipids, and a single phospholipid, ceramide, and triglyceride were identified at higher intensities in cSCC.

To the best of our knowledge, no previous molecular profiling study focused only on the lipidomic profiling of cSCC. Previous studies comparing BCC and healthy (with samples sizes of 30 and 64)[21,23] and a study comparing 12 BCC, 13 AK, and 11 healthy skin samples[22], reported lipidomic profiles for only 6 lipid groups: cholesterol, HDL, LDL, triglycerides, phospholipids, and total lipids. Triglycerides were previously reported as significantly higher in BCC vs. Healthy skin samples[22], but not significant in serum samples for the same comparison[21–23]. Phospholipids were previously found significantly higher in BCC vs. Healthy in both skin and serum samples[22].

Indeed, these studies employed lipid analysis approaches that different from used in our studies. Yet, several comparisons can still be made. For example, our study significantly improves on the level of detail of reported lipids of BCC and healthy skin. E-biopsy coupled with UPLC-MS-MS was able to measure specific ceramides, lyso-phospholipids, and diglycerides in addition to triglycerides and phospholipids. In contrast to a previous report[22], triglycerides were not identified as significant in BCC compared to Healthy skin samples. Rather, they were found in significantly lower levels in BCC compared to cSCC. Like in previous study[22], our results show higher phospholipid levels in BCC compared to Healthy skin. Six phospholipid subclasses were expressed higher in BCC compared to Healthy, and one phospholipid type was lower in BCC compared to Healthy (**Fig. 3b**).

Our study contributes novel information on the lipid profile of cSCC and its comparative analysis to BCC and to healthy skin. Diglycerides, triglycerides, ceramides, phospholipids, and lyso-phospholipids were identified in cSCC. Diglycerides were expressed significantly lower and phospholipids and lyso-phospholipids significantly higher in cSCC compared to healthy skin (**Fig. 3a**). In cSCC compared to BCC, phospholipids, lyso-phospholipids, ceramides, and triglycerides were expressed significantly higher (**Fig. 3c**). This study differs from previous cSCC studies in the methods of sample collection and lipid analysis. Previously, cSCC and healthy skin were sampled from the same patient with healthy skin taken from beyond tumor margins, and lipids were analyzed among metabolites from whole tissue samples[24]. This study, in contrast, collected healthy skin tissue from a separate set of patients and performed an exclusive lipid analysis on the collected tissues. This resulted in a larger number of lipids detected and analyzed and better reflects real life diversity of the human lipidome. Additionally, the whole tissue samples in previous studies were collected over relatively large areas, thus allowing the inherent tissue and tumor molecular heterogeneity to obscure the signals of interest. This is in contrast to localized sampling using e-biopsy that better reflects the actual spatial molecular state in the condition of interest.

The lipids included in the molecular profiling, have variable reported effects on carcinogenesis. Ceramides (Cer) are potent tumor suppressor lipids as they can enhance keratinocytes apoptosis and also block cell cycle transition limiting cancer cell proliferation[29]. Diglycosylceramides (CerG2) are involved in forming large amount of lipids in the cells of the innate immune system[29]. The higher Cer levels in cSCC compared to BCC may be a reflection of the difference in aggressiveness and the need for stronger tumor suppression in cSCC. Higher CerG2 levels in BCC compared to healthy suggest an immune response is occurring in the tumor area. Phosphatidylserine (PS) is translocated from inner to outer endothelial cell’s membrane when exposed to oxidative stress, making it a potential marker for apoptotic and tumor cells[30]. Phosphatidylcholine (PC) is involved in tumor microenvironment cellular communication and interestingly it was demonstrated that cancer cells accumulate PC precursors or products, compared to non-malignant counterparts[31]. Similarly, increased triglycerides (TG) levels were seen in both actinic keratosis and BCC compared to normal skin cells[22].

Furthermore, when diglyceride (DG or diacylglycerol), a second messenger lipid, is oxidized by UVA or UVB it may act as an endogenous tumor promoter by activation of protein kinase C (PKC) and NADPH oxidase in human neutrophils[32–35]. Lyso-phosphatidylcholine (LPC) is mainly derived from the turnover of phosphatidylcholine (PC) in the circulation by phospholipase A2 (PLA2)[36]. LPC can also induce the activation of PKC as well as phospholipase and regulate MAP kinase[36]. LPC can recruit phagocytes to the site of apoptosis, hence plays an important role in the invasion, metastasis and prognosis of tumors[36]. This aligns with our findings of higher PC and LPC in cancer groups compared to healthy skin samples. Additionally, LPC was higher in cSCC compared to BCC, potentially reflecting the more aggressive nature of cSCC. Lyso-phosphatidylinositol (LPI) is generated by PLA2 and a G protein receptor 55 (GPR55), upregulated in human cSCC and suggested to promote skin carcinogenesis and tumor aggressiveness[37]. Phosphatidylinositol (PI) derivatives are synthesized in the phosphoinositide 3-kinase (PI3K) /AKT pathway, which is one of the most frequently activated signaling pathways in human cancer, as well as been reported to be activated in both cSCC and BCC[38–40]. This supports the observed increase if PI in all comparisons groups of this study (cSCC vs. healthy, BCC vs. healthy, and cSCC vs. BCC). Lastly, Lyso-phosphatidylethanolamine (LPE)’s physiological significance in the plasma remains unknown[30], however, it has been observed to significantly increase cell proliferation in breast cancer cell lines[41].

There are few limitations in this study to be noted. Firstly, the sample size was not representative enough to draw a confident conclusion on the specific lipid behavior. Previous studies demonstrate the need for larger sample numbers to observe trends in skin cancer lipids[21,23], thus an increased sample size can improve differential expression analysis, increase the confidence in findings, and reduce the amount of falsely detected signals. Second, the study was performed on the *ex-vivo* samples, *in-vivo* sampling of skin may provide slightly different results. Third, inclusion of patients in the study was not very strict and patient lifestyle information that could provide useful information, such as levels of sun exposure, was not collected.

The e-biopsy sampling technique is still in its early stages but has the potential to be implemented in a handheld device, offering a promising solution to reduce the need for resection during biopsy. Unlike existing handheld devices like the iKnife[42,43] and MasSpec Pen[44,45], which require a real-time connection to a mass spectrometer and are primarily used intraoperatively, the e-biopsy technique shows promise for broader applications. Other needle biopsy methods such as fine-needle aspiration and core needle biopsy also eliminate the need for resection but are limited by needle size[46,47], and the need for increasing diagnostic accuracy requires more invasive procedures with larger needle diameters[48,49]. In contrast, the e-biopsy technique has demonstrated the ability to sample areas larger than an actual needle diameter[27], providing valuable site-specific information. This also stands in contrast to previous lipid analysis studies that relied on serum samples, lacking the specific spatial context. This site-specific sampling feature is advantageous as it enables mapping of tumor heterogeneity and offers a deeper understanding of tumor complexities[26].

## 5. Conclusion

This study significantly contributes to the understanding of cSCC and BCC diagnostics. It provides the first of its kind report of cSCC high throughput lipidomic profiles. It also provides high throughput lipidomic profiles for BCC and healthy skin samples, together with comparative analysis between all three tissues. Moreover, all lipidomic profiles reported here are of greater resolution compared to previous keratinocyte carcinoma lipid profiling. A total of 168 lipids were identified, of which 27 were recognized as differentially expressed in at least 1 comparison group. The differently expressed lipids are a variety of diglycerides, triglycerides, ceramides, phospholipids, and lyso-phospholipids. Overall trends indicate lower diglycerides and higher phospholipids and lyso-phospholipids in cSCC and BCC compared to Healthy skin tissue. In summary, this study significantly advances our understanding of cSCC and BCC diagnostics through high-throughput lipidomic profiling. The identification of differentially expressed lipids and the comparative analysis among the three tissue types provide crucial insights into the lipidomic alterations associated with these skin cancers. The availability of the data online and the potential of the e-biopsy technique pave the way for improved diagnostic approaches and hold promise for the future of skin cancer diagnosis.

## Data Availability

All data produced in the present work are contained in the manuscript and supplementary material.

https://github.com/GolbergLab/BCC_SCC_Lipidomics.

## Abbreviations

BCC: basal cell carcinoma
Cer: ceramides
CerG2: diglycosylceramide
cSCC: cutaneous squamous cell carcinoma
DG: diglyceride
e-biopsy: electroporation-based biopsy
LPC: lyso-phosphatidylcholine
LPE: lyso-phosphatidylethanolamine
LPI: lyso-phosphatidylinositol
PC: phosphatidylcholine
PI: phosphatidylinositol
PS: phosphatidylserine
TG: triglyceride
UPLC-MS-MS: ultra performance liquid chromatography and tandem mass spectrometry

## Data accessibility

The data that supports the findings of this study are available in the supplementary material of this article and in https://github.com/GolbergLab/BCC_SCC_Lipidomics.

## Author contributions

LL – conceptualization, experiments, lipid sampling and analysis, data preparation and analysis, bioinformatics, manuscript drafting

JW – experiments, lipid sampling and analysis

AB– experiments, samples collection, pathology, clinics, manuscript review

OS– experiments, samples collection, pathology, clinics

VK – pathology

ZY – conceptualization, data analysis

AS – conceptualization, critical manuscript review

AG– conceptualization, experiments, data analysis, manuscript drafting

EV – conceptualization, bioinformatics, manuscript drafting and approval

All authors contributed to the manuscript review.

## Acknowledgements

The authors thank Beijing Genomic Institute for lipidomics services.

## References

1. Lim HW, Collins SAB, Resneck JS, et al. The burden of skin disease in the United States. J Am Acad Dermatol. 2017;76(5):958–972.e2. 10.1016/J.JAAD.2016.12.043

2. Zhang W, Zeng W, Jiang A, et al. Global, regional and national incidence, mortality and disability-adjusted life-years of skin cancers and trend analysis from 1990 to 2019: An analysis of the Global Burden of Disease Study 2019. Cancer Med. 2021;10(14):4905–4922. 10.1002/CAM4.4046

3. Perera E, Gnaneswaran N, Staines C, Win AK, Sinclair R. Incidence and prevalence of non-melanoma skin cancer in Australia: A systematic review. Australasian Journal of Dermatology. 2015;56(4):258–267. 10.1111/AJD.12282

4. Tang E, Fung K, Chan AW. Incidence and mortality rates of keratinocyte carcinoma from 1998–2017: a population-based study of sex differences in Ontario, Canada. CMAJ. 2021;193(39):E1516–E1524. 10.1503/CMAJ.210595

5. Rogers HW, Weinstock MA, Feldman SR, Coldiron BM. Incidence Estimate of Nonmelanoma Skin Cancer (Keratinocyte Carcinomas) in the US Population, 2012. JAMA Dermatol. 2015;151(10):1081–1086. 10.1001/JAMADERMATOL.2015.1187

6. Guy GP, Ekwueme DU. Years of potential life lost and indirect costs of melanoma and non-melanoma skin cancer: A systematic review of the literature. Pharmacoeconomics. 2011;29:863–874. 10.2165/11589300-000000000-00000/FIGURES/TAB3

7. Combalia A, Carrera C. Squamous Cell Carcinoma: An Update on Diagnosis and Treatment. Dermatol Pract Concept. 2020;10(3):e2020066. 10.5826/DPC.1003A66

8. Stulberg DL, Crandell B, Fawcett RS. Diagnosis and Treatment of Basal Cell and Squamous Cell Carcinomas . Am Fam Physician. 2004;70(8):1481–1488. Accessed April 5, 2023. https://www.proquest.com/docview/234253064?pq-origsite=primo

9. Quintana RM, Dupuy AJ, Bravo A, et al. A Transposon-Based Analysis of Gene Mutations Related to Skin Cancer Development. Journal of Investigative Dermatology. 2013;133(1):239–248. 10.1038/JID.2012.245

10. Jackson SR, Jansen B, Yao Z, Ferris LK. Risk Stratification of Severely Dysplastic Nevi by Non-Invasively Obtained Gene Expression and Mutation Analyses. SKIN The Journal of Cutaneous Medicine. 2020;4(2):124–129.

11. Gerami P, Yao Z, Polsky D, et al. Development and validation of a noninvasive 2-gene molecular assay for cutaneous melanoma. Journal of the American Academy of Dermatology Volume 76, Issue 1, January 2017, Pages 114–120.e2. 2017;76(1):114–120.

12. Feingold KR, Elias PM. Role of lipids in the formation and maintenance of the cutaneous permeability barrier. Biochimica et Biophysica Acta (BBA) - Molecular and Cell Biology of Lipids. 2014;1841(3):280–294. 10.1016/J.BBALIP.2013.11.007

13. Cirenajwis H, Ekedahl H, Lauss M, et al. Molecular stratification of metastatic melanoma using gene expression profiling[]: Prediction of survival outcome and benefit from molecular targeted therapy. Oncotarget. 2015;6(14):12297–12309. www.impactjournals.com/oncotarget/

14. Priestley P, Baber J, Lolkema MP, et al. Pan-cancer whole-genome analyses of metastatic solid tumours. Nature. 2019;575(7781):210–216. 10.1038/s41586-019-1689-y

15. Malone ER, Oliva M, Sabatini PJB, Stockley TL, Siu LL. Molecular profiling for precision cancer therapies. Genome Med. 2020;12(1):1–19. 10.1186/s13073-019-0703-1

16. Vitkin E, Wise J, Berl A, et al. Proteome sampling with e-biopsy enables differentiation between cutaneous squamous cell carcinoma and basal cell carcinoma. medRxiv. Published online 2022. 10.1101/2022.12.22.22283845

17. Mun JH, Lee H, Yoon D, Kim BS, Kim MB, Kim S. Discrimination of Basal Cell Carcinoma from Normal Skin Tissue Using High-Resolution Magic Angle Spinning 1H NMR Spectroscopy. PLoS One. 2016;11(3):e0150328. 10.1371/JOURNAL.PONE.0150328

18. Huang J, Schaefer J, Wang Y, et al. Metabolic signature of eyelid basal cell carcinoma. Exp Eye Res. 2020;198:108140. 10.1016/J.EXER.2020.108140

19. Fukumoto T, Nishiumi S, Fujiwara S, Yoshida M, Nishigori C. Novel serum metabolomics-based approach by gas chromatography/triple quadrupole mass spectrometry for detection of human skin cancers: Candidate biomarkers. J Dermatol. 2017;44(11):1268–1275. 10.1111/1346-8138.13921

20. Wei L, Christensen SR, Fitzgerald ME, et al. Ultradeep sequencing differentiates patterns of skin clonal mutations associated with sun-exposure status and skin cancer burden. Sci Adv. 2021;7(1):eabd7703. 10.1126/SCIADV.ABD7703/SUPPL_FILE/ABD7703_SM.PDF

21. Anghaei S, Kamyab-Hesari K, Haddadi S, Jolehar M. New diagnostic markers in basal cell carcinoma. Journal of Oral and Maxillofacial Pathology. 2020;24(1):105. 10.4103/JOMFP.JOMFP_199_19

22. Vural P, Canbaz M, Sekçuki D, Murat A. Lipid profile in actinic keratosis and basal cell carcinoma. Int J Dermatol. 1999;38(6):439–442. 10.1046/J.1365-4362.1999.00625.X

23. Zamanian A, Rokni GR, Ansar A, Mobasher P, Jazi GA. Should variation of serum lipid levels be considered a risk factor for the development of basal cell carcinoma? Adv Biomed Res. 2014;3:108. 10.4103/2277-9175.129704

24. Chen W, Rao J, Liu Z, et al. Integrated tissue proteome and metabolome reveal key elements and regulatory pathways in cutaneous squamous cell carcinoma. J Proteomics. 2021;247. 10.1016/j.jprot.2021.104320

25. Golberg A, Sheviryov J, Solomon O, Anavy L, Yakhini Z. Molecular harvesting with electroporation for tissue profiling. Sci Rep. 2019;9(1):1–13. 10.1038/s41598-019-51634-7

26. Vitkin E, Singh A, Wise J, Ben-Elazar S, Yakhini Z, Golberg A. Nondestructive protein sampling with electroporation facilitates profiling of spatial differential protein expression in breast tumors in vivo. Scientific Reports . 2022;12(1):15835. 10.1038/s41598-022-19984-x

27. Genish I, Gabay B, Ruban A, et al. Electroporation-based proteome sampling ex vivo enables the detection of brain melanoma protein signatures in a location proximate to visible tumor margins. PLoS One. 2022;17(5):e0265866. 10.1371/JOURNAL.PONE.0265866

28. Ben-Dor A, Friedman N, Yakhini Z. Overabundance Analysis and Class Discovery in Gene Expression Data.; 2002.

29. Sugiki H, Hozumi Y, Maeshima H, Katagata Y, Mitsuhashi Y, Kondo S. C2-ceramide induces apoptosis in a human squamous cell carcinoma cell line. British Journal of Dermatology. 2000;143(6):1154–1163. 10.1046/J.1365-2133.2000.03882.X

30. Stafford JH, Thorpe PE. Increased Exposure of Phosphatidylethanolamine on the Surface of Tumor Vascular Endothelium. Neoplasia. 2011;13(4):299–IN2. 10.1593/NEO.101366

31. Saito--R--de F, Andrade--LN--de S, Bustos SO, Chammas R. Phosphatidylcholine-Derived Lipid Mediators: The Crosstalk Between Cancer Cells and Immune Cells. Front Immunol. 2022;13:768606. 10.3389/FIMMU.2022.768606/BIBTEX

32. Tanino Y, Budiyanto A, Ueda M, et al. Decrease of antioxidants and the formation of oxidized diacylglycerol in mouse skin caused by UV irradiation. Journal of Dermatological Science Supplement. 2005;1(2):S21–S28. 10.1016/J.DESCS.2005.06.003

33. Takasuka N, Takahashi M, Hori Y, et al. Promotion of mouse two-stage skin carcinogenesis by diacylglycerol-rich edible oil. Cancer Lett. 2009;275(1):150–157. 10.1016/J.CANLET.2008.10.016

34. Carsberg CJ, Ohanian J, Friedmann PS. Ultraviolet radiation stimulates a biphasic pattern of 1,2-diacylglycerol formation in cultured human melanocytes and keratinocytes by activation of phospholipases C and D. Biochemical Journal. 1995;305(2):471–477. 10.1042/BJ3050471

35. Reddig PJ, Dreckschimdt NE, Ahrens H, et al. Transgenic Mice Overexpressing Protein Kinase Cδ in the Epidermis Are Resistant to Skin Tumor Promotion by 12-O-Tetradecanoylphorbol-13-acetate. Cancer Research . 1999;59(22):5710–5718.

36. Law SH, Chan ML, Marathe GK, Parveen F, Chen CH, Ke LY. An Updated Review of Lysophosphatidylcholine Metabolism in Human Diseases. Int J Mol Sci. 2019;20(5):1149. 10.3390/IJMS20051149

37. Pérez-Gómez E, Andradas C, Flores JM, et al. The orphan receptor GPR55 drives skin carcinogenesis and is upregulated in human squamous cell carcinomas. Oncogene . 2013;32(20):2534–2542. 10.1038/onc.2012.278

38. Janus JM, O’Shaughnessy Rfl, Harwood CA, Maffucci T. Phosphoinositide 3-Kinase-Dependent Signalling Pathways in Cutaneous Squamous Cell Carcinomas. Cancers . 2017;9(7):86. 10.3390/CANCERS9070086

39. Hafner C, Landthaler M, Vogt T. Activation of the PI3K/AKT signalling pathway in non-melanoma skin cancer is not mediated by oncogenic PIK3CA and AKT1 hotspot mutations. Exp Dermatol. 2010;19(8):e222–e227. 10.1111/J.1600-0625.2009.01056.X

40. Liu P, Cheng H, Roberts TM, Zhao JJ. Targeting the phosphoinositide 3-kinase pathway in cancer. Nature Reviews Drug Discovery . 2009;8(8):627–644. 10.1038/nrd2926

41. Park SJ, Lee KP, Im DS. Action and Signaling of Lysophosphatidylethanolamine in MDA-MB-231 Breast Cancer Cells. Biomol Ther (Seoul). 2014;22(2):135. 10.4062/BIOMOLTHER.2013.110

42. Balog J, Sasi-Szabó L, Kinross J, et al. Intraoperative Tissue Identification Using Rapid Evaporative Ionization Mass Spectrometry.; 2013. https://www.science.org

43. Tzafetas M, Mitra A, Paraskevaidi M, et al. The intelligent knife (iKnife) and its intraoperative diagnostic advantage for the treatment of cervical disease. Proc Natl Acad Sci U S A. 2020;117(13):7338–7346. 10.1073/PNAS.1916960117/SUPPL_FILE/PNAS.1916960117.SAPP.PDF

44. Sans M, Zhang J, Lin JQ, et al. Performance of the MasSpec pen for rapid diagnosis of Ovarian cancer. Clin Chem. 2019;65(5):674–683. 10.1373/clinchem.2018.299289

45. Zhang J, Rector J, Lin JQ, et al. Nondestructive Tissue Analysis for Ex Vivo and in Vivo Cancer Diagnosis Using a Handheld Mass Spectrometry System.; 2017. https://www.science.org

46. Helbich TH, Rudas M, Haitel A, et al. Evaluation of needle size for breast biopsy: comparison of 14-, 16-, and 18-gauge biopsy needles. American Journal of Roentgenology. 1998;171(1):59–63. 10.2214/AJR.171.1.9648764

47. Serefoglu EC, Altinova S, Ugras NS, Akincioglu E, Asil E, Balbay D. How reliable is 12-core prostate biopsy procedure in the detection of prostate cancer? Canadian Urological Association Journal. 2013;7(5-6):e293–8. 10.5489/cuaj.1248

48. Shah VI, Raju U, Chitale D, Deshpande V, Gregory N, Strand V. False-negative core needle biopsies of the breast. Cancer. 2003;97(8):1824–1831. 10.1002/CNCR.11278

49. Rodrigues LKE, Leong SPL, Ljung BM, et al. Fine needle aspiration in the diagnosis of metastatic melanoma. J Am Acad Dermatol. 2000;42(5I):735–740. 10.1067/mjd.2000.103812

