## Supplementary figures and images for "Comparison of lipidomic profiles sampled with electroporation-based biopsy from healthy skin, squamous cell carcinoma, and basal cell carcinoma"

### Fig. S1

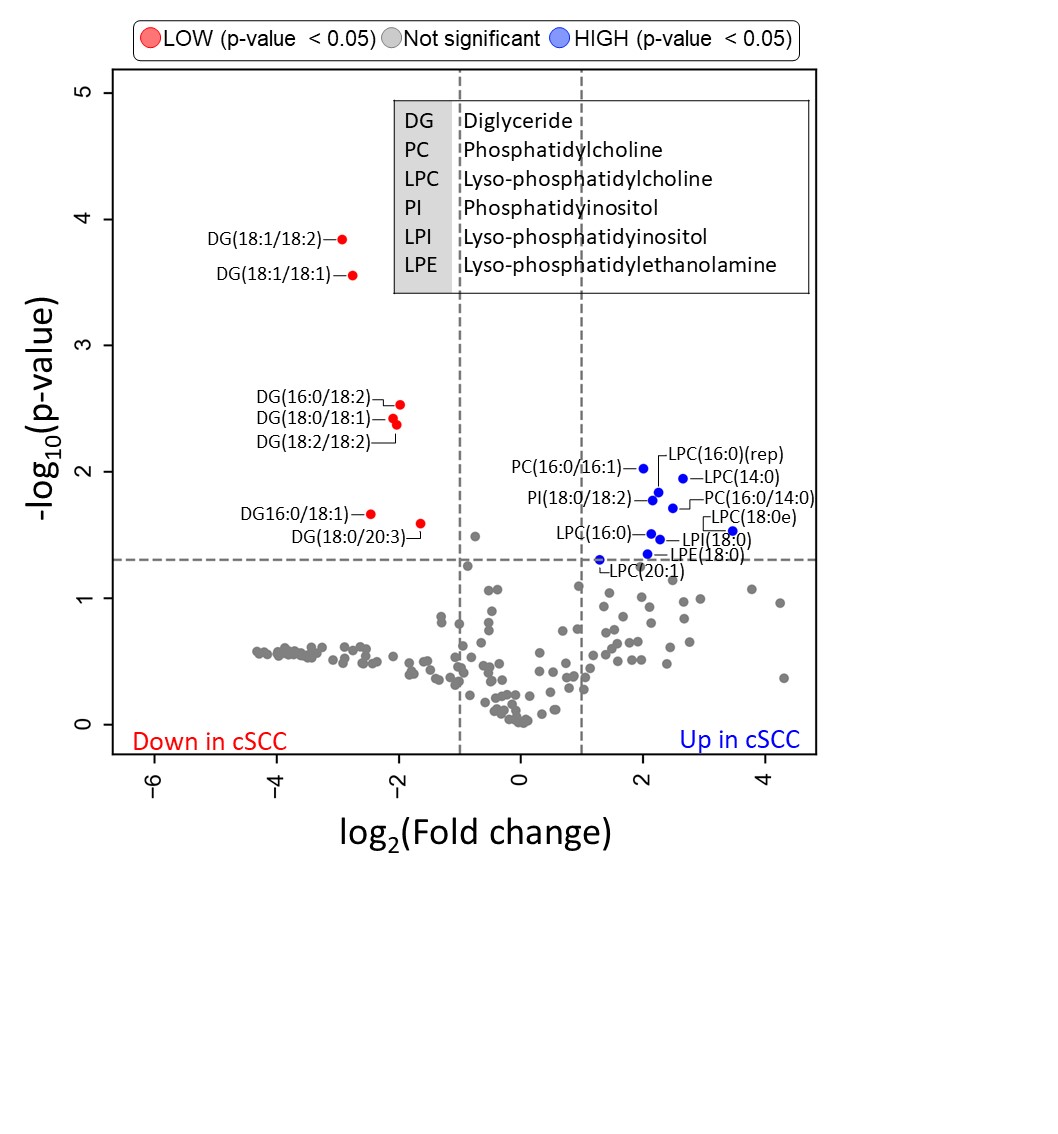

### Fig. S2

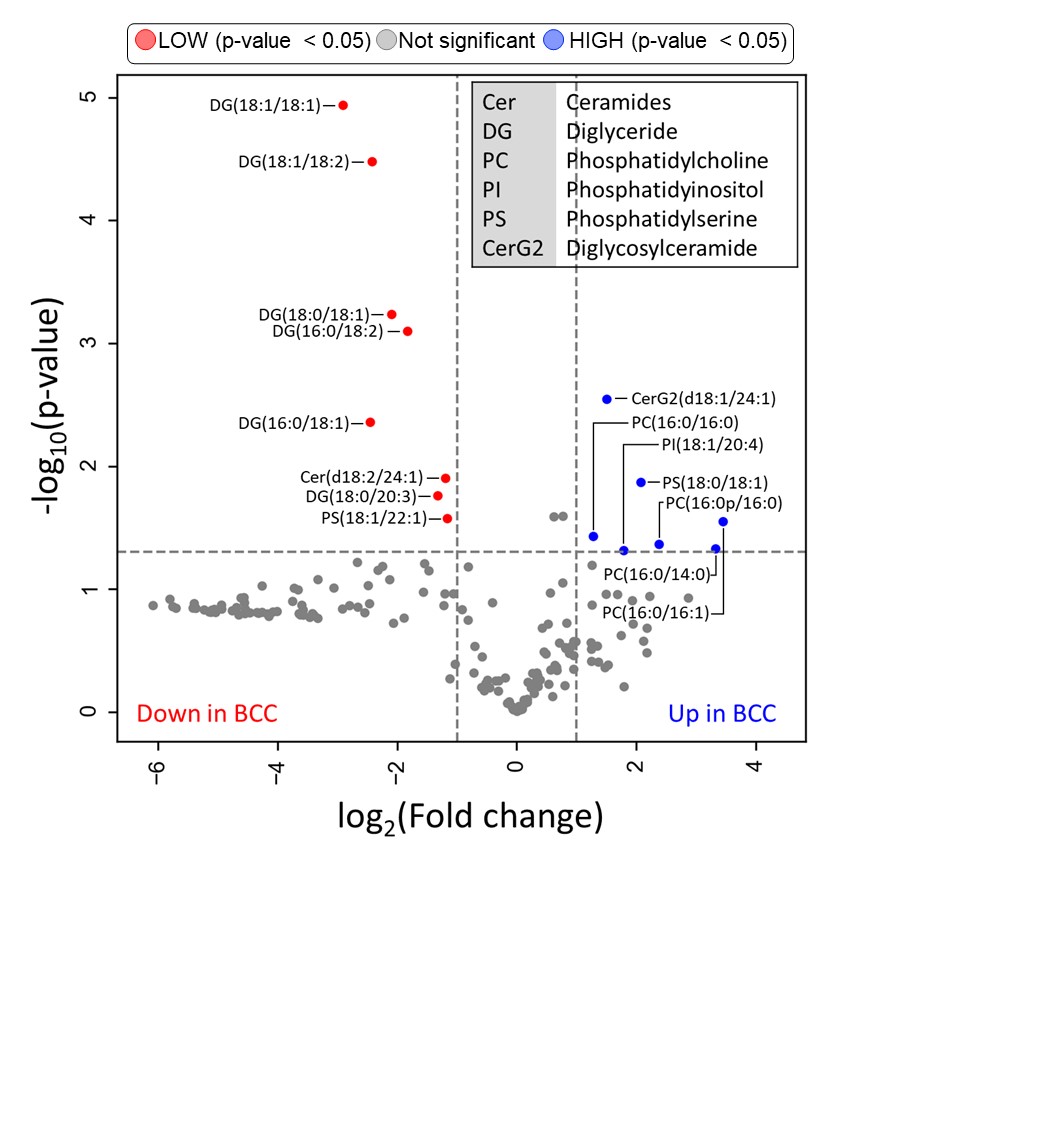

### Fig. S3

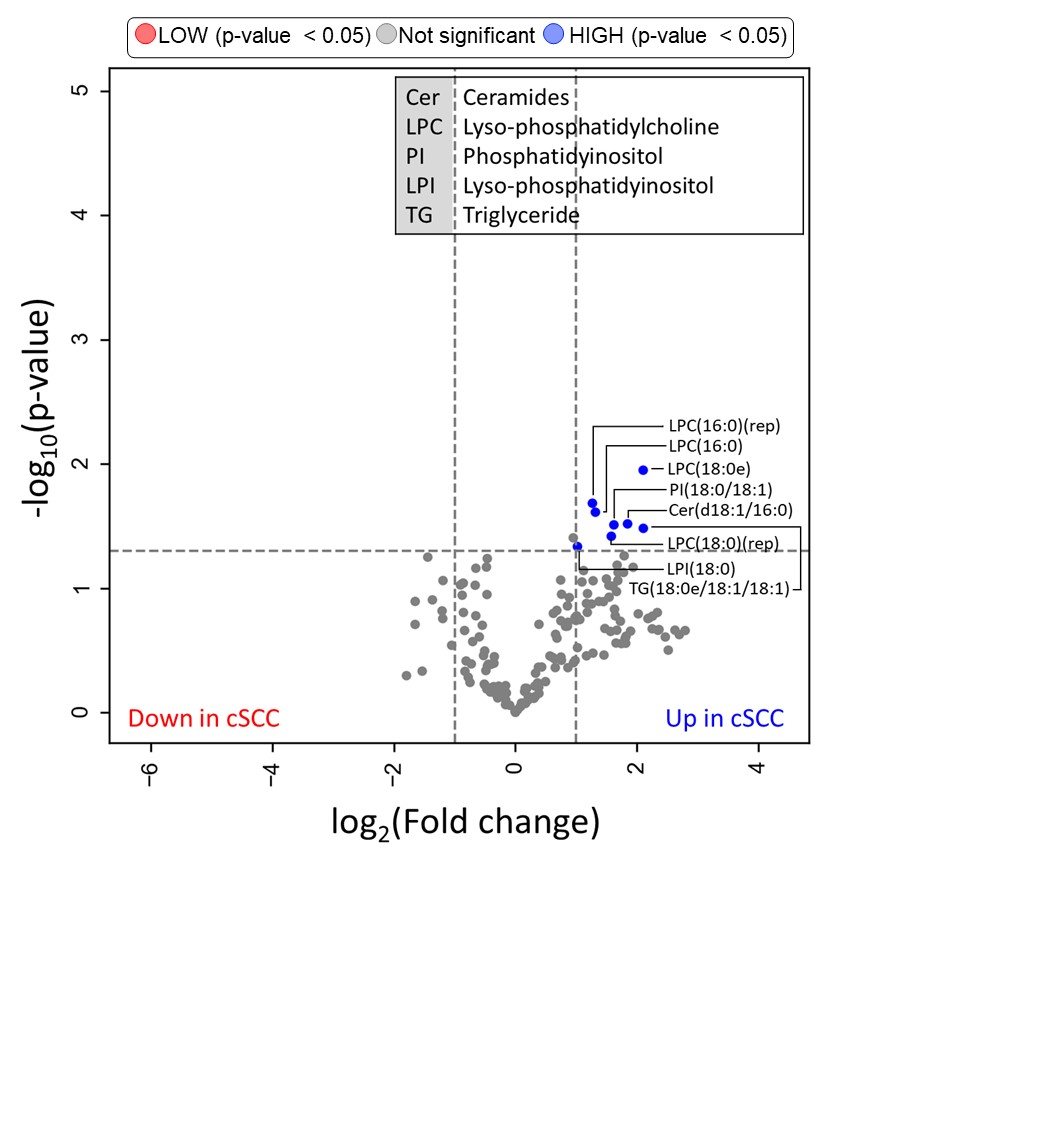
