## Supplementary Methods for "Comparison of lipidomic profiles sampled with electroporation-based biopsy from healthy skin, squamous cell carcinoma, and basal cell carcinoma"

### UPLC-MS-MS Analysis

The following sections are presented as reported by BGI Group.

#### Main Instruments and Reagents

Ultra high-performance liquid phase (Waters 2D UPLC, Waters, USA)

High resolution mass spectrometer (Q Exactive, Thermo Fisher Scientific, USA)

The chromatographic columns:

ACQUITY UPLC CSH C18(1.7 μm,2.1*100 mm, Waters, USA)

ACQUITY UPLC BEH C18(1.7 μm,2.1*100 mm, Waters, USA)

ACQUITY UPLC BEH Amide (1.7 μm,2.1*100 mm, Waters, USA)

Low temperature high speed centrifuge (Centrifuge 5430, Eppendorf)

Vortex (QL-901, Kylin-Bell Lab Instruments Co.,Ltd., China)

Milli-Q apparatus (Milli-Q Integral,Millipore Corporation, USA)

Freezing vacuum concentrator (Maxi Vacbeta, GENE COMPANY）

SPLASH Internal standards: (330707, SPLASHTM Lipidomix Mass Spec Standard, Avanti Polar Lipids, USA), the stock solution concentration of each lipid standard is as follows: LPC 18:1(d7), 25 μg/mL; LPE 18:1(d7), 5 μg/mL; PC 15:0–18:1(d7), 160 μg/mL; PE 15:0–18:1(d7), 5 μg/mL; PG 15:0–18:1(d7), 30 μg/mL; PS 15:0–18:1(d7), 5 μg/mL; PI 15:0–18:1(d7), 10 μg/mL; PA 15:0–18:1(d7), 7 μg/mL; SM d18:1–18:1(d9), 30 μg/mL; cholesterol(d7), 100 μg/mL; CE 18:1(d7), 350 μg/mL; MG 18:1(d7), 2 μg/mL; DG 15:0–18:1(d7), 10μg/mL; and TG 15:0–18:1(d7)–15:0, 55 μg/mL

Methanol（A454-4) and Acetonitrile (A996-4) and Isopropanol (A461-4) were of LCMS grade (Thermo Fisher Scientific, USA); Ammonium formate (17843-250G, Honeywell Fluka, USA), Formic acid (50144-50ml, DIMKA, USA) and water was purified by a Milli-Q apparatus.

#### Lipids Extraction

Samples were thawed at 4°C. 100 μL of supernatant was transferred to a 96-well microplate. Next, 300 μL of precooled (-20°C) isopropanol and 10 μL of SPLASH internal standards solution were added. The mixture was subsequently vortexed and stored overnight at -20°C. The samples were then centrifuged at 4°C for 20 minutes at 4000 rpm. Supernatant was transferred to 1.5 mL vial. From each sample, 10 μL was mixed with QC samples for evaluation of repeatability and stability of the LC-MS analysis.

Lipids separation and detection was performed on a LC-MS system using Waters 2D UPLC (Waters, USA) and Q Exactive high resolution mass spectrometer (Thermo Fisher Scientific, USA).

### Lipids UPLC-MS-MS Analysis

Liquid chromatography conditions: A CSH C18 column (1.7 μm 2.1*100 mm, Waters, USA) was used. The mobile phase consisted of solvent A (60% acetonitrile aqueous solution + 0.1% formic acid + 10 mM ammonium formate) and solvent B (10% acetonitrile aqueous solution + 90% Isopropanol + 0.1% formic acid + 10 mM ammonium formate) under positive ion mode, and solvent A (60% acetonitrile aqueous solution + 10 mM ammonium formate) and solvent B (10% acetonitrile aqueous solution + 90% Isopropanol + 10 mM ammonium formate) under negative ion mode. Gradient elution conditions were set as follows: 0~2 min, 40% to 43% B; 2~2.1 min, 43% to 50% B; 2.1~7 min, 50% to 54% B; 7~7.1 min, 54% to 70% B; 7.1~13 min, 70% to 99% B; 13~13.1 min, 99% to 40% B; 13.1~15 min, 40% B. The flow rate was 0.35 mL/min. The column oven was maintained at 55°C. The injection volume was 5 μL.

Mass spectrometry conditions: A Q Exactive mass spectrometer (Thermo Fisher Scientific, USA) was used to obtain MS1 and MS2 data. The MS scan method was in the range of m/z 200–2000, The MS1 resolution was 70,000, AGC was 3e6, and the maximum injection time was 100 ms. According to the precursor ion intensity, the top 3 ions were selected for MS2 analysis, MS2 resolution was 17,500, AGC was 1e5, maximum injection time was 50 ms, and collision energy (stepped and NCE) were set as: 15, 30 and 45 eV. The parameters of ESI were: sheath gas of 40 L/min, aux gas of 10 L/min, spray voltage(|KV|) of 3.80 in positive ion mode and of 3.20 in negative ion mode, capillary temperature of 320°C and aux gas heater temperature of 350°C.

In order to provide more reliable experimental results during instrument detection, random sorting of samples was carried out to reduce systematic errors. Every 10 samples are interspersed with one QC sample for testing.

### Lipids Data Preprocessing

The raw file generated by LC-MS/MS detection was imported into LipidSearch v.4.1 software (Thermo Fisher Scientific, USA)[1] for lipid molecular identification and quantification with following parameters: product as the identification type; 5ppm as the quality deviation threshold between precursor ion and product ion in library; 5.0% as threshold for relative response deviation from product ion; 5 ppm as the mass deviation of peak extraction; M-score was set as 5.0, c-score was set as 2.0, and the identification level was selected as {"A", "B", "C" ,"D"}. All lipid categories were selected for identification; Adduct forms of positive ion mode were [M+H]+, [M+NH4]+, [M+Na]+, and of negative ion mode were [M-H]- and [M-2H]- and [M-HCOO]-.

All identified lipids were peak-aligned, and not-rejected results were considered for further analysis. The peak alignment method was set as Mean, the retention time deviation was set as 0.1 min, the peak filtering was set as New Filter, top rank, all isomer peak, and the identification level were selected as "A: Lipid categories and all fatty acid chains can be completely identified", "B: Category-specific ions and fatty acid fragment ions can be detected", "C: Class-specific ions or fatty acid fragment ions, only one of which can be detected", and "D: Lipid structures, such as dehydrated ions, cannot be recognized".

The lipids identified and quantified by LipidSearch were imported into metaX[2] for data preprocessing and subsequent analysis. The data was first filtered to remove lipid molecules missing more than 50% of QC samples and more than 80% of experimental samples. Second, K-nearest Neighbor algorithm (KNN) was used to fill missing lipid values in the remaining samples with weighted average peak intensity using calculations of Euclidean Distance. Third, Probabilistic Quotient Normalization (PQN) was used to normalize the data and obtain relative peak area. Finally, the data was further filtered to remove lipid molecules whose Coefficient of Variation (CV) of relative peak area was greater than 30% in all QC samples.

### Data Quality Control

The data quality was evaluated by the repeatability of QC sample detection. Quality control consisted of overlapping QC chromatograms to ensure signal stability through minimal fluctuation of retention time and peak response intensity. Principal component analysis (PCA) of log2 transformed data standardized with pareto scaling was used to observe separation trends between sample groups, identify abnormal points, and variation between and within groups. In this way, the overall distribution and stability of QC samples and all samples can be observed. Lastly, the differences in the number and peak area of lipid molecules underwent QC by examining the ratio of lipid molecules whose CV of relative peak area is less than or equal to 30% in QC samples to the number of all detected compounds. Data was qualified in the ratio was greater or equal to 60%.

### References

1. Chauhan MZ, Valencia AK, Piqueras MC, Enriquez-Algeciras M, Bhattacharya SK. Optic nerve lipidomics reveal impaired glucosylsphingosine lipids pathway in glaucoma. *Invest Ophthalmol Vis Sci*. 2019;60(5):1789-1798. doi:10.1167/iovs.18-25802

2. Wen B, Mei Z, Zeng C, Liu S. metaX: A flexible and comprehensive software for processing metabolomics data. *BMC Bioinformatics*. 2017;18:1-14. doi:10.1186/s12859-017-1579-y
